# Longitudinal Evolution of Diffusion Metrices in the Cerebello-Thalamo-Cortical Tract After MRgFUS Thalamotomy for Essential Tremor

**DOI:** 10.1101/2024.12.06.24318287

**Authors:** Neeraj Upadhyay, Marcel Daamen, Veronika Purrer, Valeri Borger, Carsten Schmeel, Jonas Krauß, Angelika Maurer, Alexander Radbruch, Ullrich Wüllner, Henning Boecker

## Abstract

Magnetic resonance-guided focused ultrasound (MRgFUS) thalamotomy in essential tremor (ET) targets the ventral intermediate nucleus hub region within the cerebello-thalamo-cortical tract (CTCT). Understanding the microstructural changes in the CTCT over time and their link to tremor improvement is crucial from a tremor-network perspective. We retrospectively analyzed tremor scores, lesion characteristics, and diffusion MRI-derived CTCT microstructural measures in 27 ET patient’s pre-treatment (T0), at 1 month (T2), and 6 months (T3) post-MRgFUS. Using probabilistic tractography, we created an average CTCT mask for assessing fractional anisotropy (FA), axial (AD), mean (MD), and radial diffusivity (RD) measures across time points. Significant tremor reduction was observed at T2 and T3. The Linear mixed effect analyses showed significant time effects for FA, MD, and AD. Relative to baseline, post-hoc comparisons showed a significant decrease of FA and AD at lesion site only for T2. Instead, there was a significant increase in AD and MD at T3 compared to T2 at lesion site, and remotely near the motor cortex. Lesion size and FA changes in the CTCT at T2 showed only trend-level correlations with tremor outcome. Stronger associations were observed for the thalamic lesion-tract overlap at T2, which were even more robust at T3. Dynamic microstructural changes suggest early axonal disruption at the lesion site and subsequent reorganization, with remote CTCT changes potentially indicating chronic degeneration. Meanwhile, microstructural measures show limited predictive value for longer-term tremor outcome compared with macroanatomical lesion-CTCT overlap. Yet, advanced diffusion imaging protocol could increase the sensitivity to predict MRgFUS clinical outcome.

## INTRODUCTION

Essential tremor (ET) is a common movement disorder (Elble, 2013) and has recently been recognized as the most common form of cerebellar degeneration (Louis & Faust, 2020). The pathophysiology of ET involves rhythmic oscillations that propagate from the cerebellum via the ventral intermediate nucleus (VIM) of the thalamus along the cerebello-thalamo-cortical tract (CTCT) to motor cortical areas (Helmich et al., 2013; Lenka et al., 2017; Welton et al., 2021).

Neuromodulation methods like deep brain stimulation (DBS) and magnetic resonance-guided focused ultrasound (MRgFUS) (Germann et al., 2024; Giordano et al., 2020) have been reported as effective treatments to alleviate drug-resistant tremor in ET (Agrawal et al., 2021; Lak et al., 2022; W. K. Miller et al., 2022). The VIM serves as a key therapeutic target for both neuromodulation methods in ET (Gallay et al., 2008). In particular, MRgFUS creates a permanent focal brain lesion in the VIM by applying thermal ablation via focused ultrasound (Haar & Coussios, 2007). The localized tissue damage at the lesion may trigger chronic fiber degeneration and/or fiber rearrangements within the CTCT, both locally and distantly from the lesion (Wintermark, Huss, et al., 2014; Pineda-Pardo et al., 2019; Zur et al., 2020; Mazerolle et al., 2021; Thaler et al., 2023). Understanding the dynamics of microstructural alterations in the CTCT resulting from MRgFUS thalamotomy is essential to determine their impact on treatment outcomes and the nature of any ensuing degeneration and reorganization.

The amount of overlap of the lesion extent with the CTCT tract is suggested to be related to clinical tremor improvement (Chazen et al., 2018; Purrer et al., 2021). Beyond lesion volume, advanced neuroimaging techniques, such as diffusion-weighted imaging (DWI), have been used to target specific tracts in ET and to assess white matter (WM) microstructural changes due to MRgFUS thalamotomy (Hori et al., 2020; Purrer et al., 2023). Decreased fractional anisotropy (FA) and increased mean diffusivity (MD) provide information about the loss of microstructural integrity (Tae et al., 2018). Axial diffusivity (AD) and radial diffusivity (RD) can provide additional information on axonal damage or demyelination, respectively (Song et al., 2005; Sun et al., 2008). While the specificity of these measures in identifying pathology is still debated (Jones et al., 2013), AD and RD provide sensitive indicators for microstructural alterations after treatment (Winston et al., 2014). Recent studies have reported microstructural alterations in the CTCT associated with tremor reduction (Chazen et al., 2018; Miller et al., 2022; Thaler et al., 2023). Relative local decreases in FA in the targeted VIM have been linked to tremor improvement (Wintermark, Huss, et al., 2014; Hori et al., 2020; Zur et al., 2020; Thaler et al., 2023) within a 3- to 6-month duration following MRgFUS in ET. Moreover, local and distant increases in AD and RD along the CTCT have been reported over follow-up periods of three months (Pineda-Pardo et al., 2019; Mazerolle et al., 2021). Even one year after MRgFUS thalamotomy, decreases in FA and increases in MD and RD were reported, both locally and remotely (Thaler et al., 2023). These studies shed light on potential underlying mechanisms of tract reorganization, notably axonal damage or reorganization and demyelination.

However, these studies frequently have relatively small sample sizes (Wintermark, Huss, et al., 2014; Hori et al., 2020; Thaler et al., 2023). Regarding the clinical significance of the observed changes, existing studies also demonstrate inconsistencies when correlating acute microstructural changes (1-3 days post-MRgFUS) with long-term clinical outcomes (Pineda-Pardo et al., 2019; Hori et al., 2020; Thaler et al., 2023; Zur et al., 2020). These are possibly due to the influence of local edema, compromising the specificity of the microstructural measures. Therefore, it is crucial to assess the relationship between microstructural measures and clinical outcomes when edema is largely regressed to gain more accurate insights. Finally, the intermediate evolution of diffusion metrics following MRgFUS lesioning is limited by the fact that many publications to date either do not include DWI measurements after three months of intervention (Wintermark, Huss, et al., 2014; Pineda-Pardo et al., 2019; Mazerolle et al., 2021) or, on the other hand, only focus on long-term outcomes after one year (Hori et al., 2020; Thaler et al., 2023).

In the present study, we examined a comparatively large cohort of ET patients including intermediate time points i.e., before, one month, and six months after MRgFUS thalamotomy, to assess both, short-term and long-term intervention effects on voxel-wise DWI measures of microstructural integrity within the CTCT, both in the treated and untreated (for control) hemisphere. We hypothesized that 1) while microstructural changes occur at the lesion site are still detectable after short-term recovery following MRgFUS, additional remote changes along the CTCT tract will only manifest during long-term follow-up. Moreover, we assessed 2) whether the observed lesion-specific and remote alterations in DWI microstructural measures were associated with the amount of tremor reduction at one month and six months, respectively. For comparison, we also 3) examined the relationship of clinical outcome with two macroanatomical imaging measures: the overall lesion volume and the overlap between the lesion and CTCT.

## METHODS

Twenty-seven patients (23 right-hand and 4 left-hand dominant) with severe drug-refractory ET (defined as at least two insufficient prior medication treatments) received unilateral MRgFUS treatment between October 2018 and October 2023. According to the MDS consensus criteria (Bhatia et al., 2018), a neurologist with expertise in movement disorders verified the ET diagnosis. Exclusion criteria included brain injury, epilepsy, coagulopathies, severe heart disease, a history of psychiatric illnesses or substance abuse, cognitive impairment, and a skull density ratio below 0.3, as this influences the MRgFUS treatment accuracy. The details of inclusion criteria for MRgFUS are provided in the German Clinical Trials registry (DRKS00016695).

The Institutional Review Board (Ethikkommission an der Medizinischen Fakultät Bonn) gave their approval (Ethics no. 314/18), and the study was carried out in accordance with the Declaration of Helsinki. Every Participant provided informed written consent before inclusion.

The detailed protocol for MRgFUS treatment has been described previously (Keil et al., 2020; Purrer et al., 2021; Pohl et al., 2022). In brief, the patient’s head was fixed in a stereotactic frame in the focused ultrasound system (ExAblate 4000 System, InSightec, Haifa, Israel) while the procedure was carried out in a 3-Tesla (3T) MRI system (Discovery MR750w, GE Healthcare, Chicago, IL, USA). The thalamic VIM contralateral to the hand most affected by tremor was chosen as target. The standard stereotactic coordinates used for the VIM were 14 mm lateral to the midline or 11 mm lateral to the wall of the third ventricle and 25% anterior to the posterior commissure along the intercommissural line. Subtle adaptations of the target coordinates were made according to intraoperative clinical assessments regarding optimal tremor reductions, side effects, and pre-treatment DTI assessments of CTCT coordinates, as well as medial lemniscus and corticospinal tracts. Finally, a permanent lesion was produced (temperature range of 55 to 60°C) using multiple sonications at sufficient energy levels.

### Clinical evaluation

Clinical evaluations were carried out by two trained neurologists two days before (T0), as well as three days (T1), one month (T2), and six months (T3) after MRgFUS treatment. The assessment included the Fahn and Tolosa Clinical Rating Scale for Tremor (CRST) (Fahn et al., 1988), which is frequently used as a severity score in the clinical evaluation of ET (Elble et al., 2013): A modified composite (CRST-AB) including seven items was assembled from its subscale B and the upper extremity tremor items of subscale A. Greater tremor severity was indicated by higher CRST-AB scores (maximum: 28). We calculated separate CRST-AB scores for both, the treated and the untreated side.

### MRI acquisition

MRI was obtained at T0, T1, T2, and T3 following MRgFUS therapy using a 3T scanner (Philips Achieva TX, Best, The Netherlands) equipped with an eight-channel head coil. The protocol included T1-weighted magnetization-prepared rapid gradient echo (MPRAGE) sequences (1 mm isotropic resolution; TR = 7.286 ms, TE = 3.93 ms, matrix = 256 x 256 mm, total scan time = 279.6 sec), as well as DWI acquisitions using single-shot echo-planar spin-echo sequences with following parameters: 70 axial 2-mm thick slices, no gap, and 56 gradient directions (TR = 7.78 s, TE = 65 ms, matrix = 112 x 112 mm, six images with b=0 and rest b= 1200 sec/mm^2^).

### MR data processing

#### Lesion volume segmentation

Due to the presence of local edema, which is a temporary phenomenon (Keil et al., 2020), we didn’t use the data immediately following MRgFUS treatment (T1) in our analysis. First, T1-weighted images were used to assess the lesion volume at T2 and T3. One rater (VP) manually delineated the lesions in each individual at each time point using the 3D editing tool in ITK-SNAP (version 3.6.0; https://www.itk-snap.org/) and calculated the lesion volume by obtaining their 3D native space volumes in mm^3^.

#### Lesion-CTCT overlap

To derive a tract-specific estimate of lesion extent, the individual overlap was calculated between lesions from T2 and T3 and CTCT. For this purpose, the manually segmented lesions at T2 and T3 were transformed to T0. The overlap between the CTCT and the binary lesion was calculated using the following formula <u>(Ghielmetti et al., 2024)</u>:

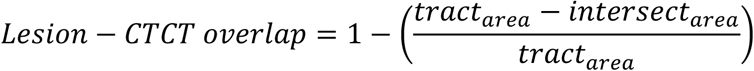

where intersect_area_ is the overlap between the tract and the lesion in the same slice, and tract_area_ is the area of the tract that corresponds to the maximal extent of the lesion, represented in the number of pixels.

#### Microstructural parameters

DWI data from T0, T2, and T3 were analyzed using FDT toolbox (https://fsl.fmrib.ox.ac.uk/fsl/fslwiki/FDT) available in FSL-6.04. The preprocessing pipeline included susceptibility distortion, motion, and eddy current correction. Susceptibility distortion correction was carried out using the opposite phase encoding non-diffusion (b0) image by applying the *top-up* tool. Later, the susceptibility distortion parameters were inserted in the *eddy* tool in order to correct for susceptibility, motion, and eddy currents in a single step. This step also produced the vector direction after correcting for rotation. The eddy current-corrected diffusion data and the rotated vectors were used to estimate the fiber orientation using the *bedpostX* tool. The output was used to perform the probabilistic tractography.

Probabilistic tractography was performed to estimate the location of the CTCT using the ipsilateral dentate nucleus as seed region and passing through the ipsilateral superior cerebellar peduncle (SCP), the contralateral thalamus and the contralateral precentral gyrus as waypoints in reference to the treated side. Seeds and waypoints were extracted from standard atlases: dentate nucleus (He et al., 2017), SCP (ICBM-DTI-81 white-matter labels atlas (Mori et al., 2006), thalamus (individual space parcellation using the model-based segmentation tool FSL first; https://fsl.fmrib.ox.ac.uk/fsl/fslwiki/FIRST), and precentral gyrus (Harvard-Oxford cortical atlas in the MNI 1 mm space). All the atlas-based regions of interest were nonlinearly registered to the individual space. Later, a linear rigid transformation was applied to bring the seed and waypoints into diffusion space. For tractography, 5000 streamlines were generated from the seed region using the standard parameters of the Probtracx2 function in the FDT toolbox. To construct the diffusion metrics, we used the weighted least square fitting approach and created FA, AD, RD, and mean diffusivity (MD) maps to evaluate the microstructural changes from T0 to T2, T0 to T3 and T2 to T3. In three subjects, the diffusion maps were flipped from right to left to bring the MRgFUS thalamotomy sites to the same side and to allow group statistical comparisons. Later, the respective FA, AD, RD, and MD maps were nonlinearly registered to the MNI ICBM 152 non-linear 6th Generation Symmetric atlas, available in FSL (Grabner et al., 2006). Further, we implemented a 5mm full width half maximum (FWHM) Gaussian kernel to smooth the data.

### Statistics

Clinical scores, lesion volumes and lesion-CTCT overlap were analyzed using Jamovi (version: 2.3.28.0; https://www.jamovi.org). We performed a repeated measures ANOVA to compare CRST-AB scores at different time points for both, treated and nontreated sides. For lesion volume and lesion-CTCT overlap pairwise comparisons between T2 and T3 were performed. Significant results were reported at p<0.05, after implementing the multiple comparison correction for post-hoc testing. Effect sizes were reported as eta squared (η^2^) value for the F-test and as Cohen’s d (d) for the t-statistics.

To assess the microstructural changes within the CTCT over time, we performed linear mixed effect (LME) modeling using the 3dLmer tool from AFNI (https://afni.nimh.nih.gov/pub/dist/doc/program_help/3dLMEr.html). A simple model was created including time as a fixed effect and subjects as a random effect. This produces main effects of time and post-hoc pairwise comparisons between implemented contrast to assess the direction of changes between T0, T2, and T3. Based on a cluster-defining height threshold of p<0.001 voxelwise, we implemented multiple comparison correction using the updated ACF approach to calculate the amount of smoothing and applied a simulation approach to identify clusters surviving family-wise error (FWE) correction at α < 0.05 with corresponding cluster size (k). Furthermore, we corrected the post-hoc comparisons by adjusting for the number of comparisons (α=0.0166 (0.05/3)) and represented results with corresponding cluster sizes.

To explore the clinical relevance of the observed microstructural changes after lesioning, we performed correlation analyses between the percent change in CRST-AB tremor scores and imaging parameters, including (A) global lesion volume (Nkomboni et al., 2024) and (B) the lesion-CTCT overlap, which were already examined in earlier studies (Kapadia et al., 2020), as well as (C) the relative diffusion metric changes obtained from significant clusters in our DWI analyses. We employed Kendall’s tau-b (τ) rank-based correlation (Kendall, 1938) due to its higher accuracy in calculating p-values for smaller sample sizes and its robustness against data discrepancies (Arndt et al., 1999). Significant correlations were reported at p < 0.05.

## RESULTS

### Clinical scores

All demographic and clinical measures are provided in Table 1. Repeated measures ANOVA for the treated side showed significant changes in CRST AB subscale over time after correcting for sphericity using the Greenhouse-Geisser method (F(1.6)=154, p <0.001, η^2^=0.682) (Figure 1A). Post-hoc comparisons indicated a significant reduction for CRST-AB at T2 (M±SD = 5.07 ± 4.27; t(26) = −13.7, p <0.001, d = −2.649) compared to T0 (M±SD = 18.78 ± 4.27). The CRST-AB remained reduced at T3 relative to T0 (M±SD = 5.85 ± 4.56; t(26) = −13.43, p <0.001, d = - 2.585). No significant differences were observed in CRST-AB between T2 and T3 (t(26) = −1.25, p =0.437, d = −0.240). Repeated measures ANOVA for the untreated side showed no significant changes in CRST-AB over the time (F(2)=0.316, p =0.730, η^2^=0.002) (Figure 1B).

**Table 1.**
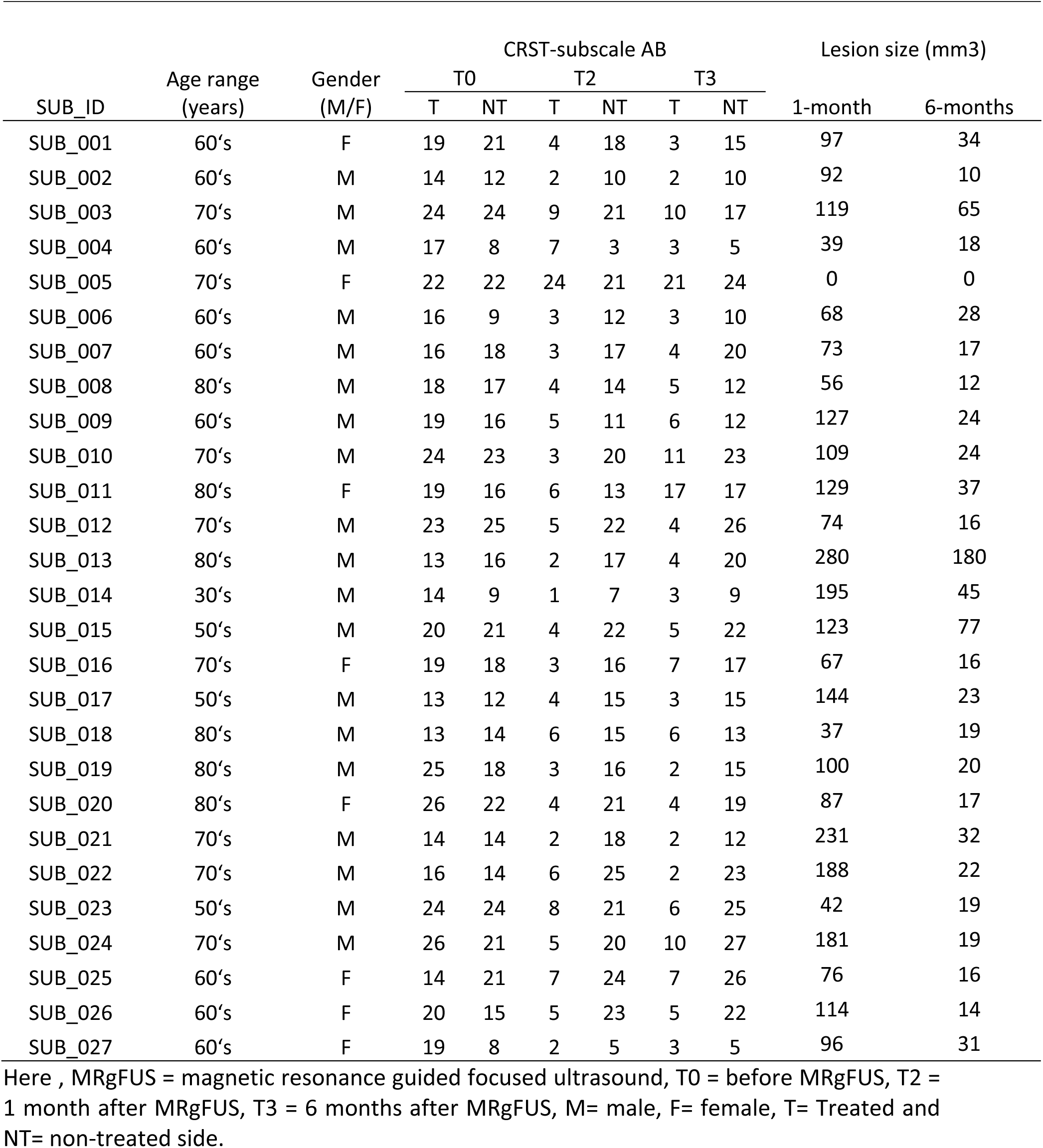
Clinical and demographic data of 27 Essential Tremor patients.

| SUB_ID | Age range<br>(years) | Gender<br>(M/F) | CRST-subscale AB |  |  |  |  |  | Lesion size (mm3) |  |
| --- | --- | --- | --- | --- | --- | --- | --- | --- | --- | --- |
|  |  |  | T0 |  | T2 |  | T3 |  | 1-month | 6-months |
|  |  |  | T | NT | T | NT | T | NT |  |  |
| SUB_001 | 60's | F | 19 | 21 | 4 | 18 | 3 | 15 | 97 | 34 |
| SUB_002 | 60's | M | 14 | 12 | 2 | 10 | 2 | 10 | 92 | 10 |
| SUB_003 | 70's | M | 24 | 24 | 9 | 21 | 10 | 17 | 119 | 65 |
| SUB_004 | 60's | M | 17 | 8 | 7 | 3 | 3 | 5 | 39 | 18 |
| SUB_005 | 70's | F | 22 | 22 | 24 | 21 | 21 | 24 | 0 | 0 |
| SUB_006 | 60's | M | 16 | 9 | 3 | 12 | 3 | 10 | 68 | 28 |
| SUB_007 | 60's | M | 16 | 18 | 3 | 17 | 4 | 20 | 73 | 17 |
| SUB_008 | 80's | M | 18 | 17 | 4 | 14 | 5 | 12 | 56 | 12 |
| SUB_009 | 60's | M | 19 | 16 | 5 | 11 | 6 | 12 | 127 | 24 |
| SUB_010 | 70's | M | 24 | 23 | 3 | 20 | 11 | 23 | 109 | 24 |
| SUB_011 | 80's | F | 19 | 16 | 6 | 13 | 17 | 17 | 129 | 37 |
| SUB_012 | 70's | M | 23 | 25 | 5 | 22 | 4 | 26 | 74 | 16 |
| SUB_013 | 80's | M | 13 | 16 | 2 | 17 | 4 | 20 | 280 | 180 |
| SUB_014 | 30's | M | 14 | 9 | 1 | 7 | 3 | 9 | 195 | 45 |
| SUB_015 | 50's | M | 20 | 21 | 4 | 22 | 5 | 22 | 123 | 77 |
| SUB_016 | 70's | F | 19 | 18 | 3 | 16 | 7 | 17 | 67 | 16 |
| SUB_017 | 50's | M | 13 | 12 | 4 | 15 | 3 | 15 | 144 | 23 |
| SUB_018 | 80's | M | 13 | 14 | 6 | 15 | 6 | 13 | 37 | 19 |
| SUB_019 | 80's | M | 25 | 18 | 3 | 16 | 2 | 15 | 100 | 20 |
| SUB_020 | 80's | F | 26 | 22 | 4 | 21 | 4 | 19 | 87 | 17 |
| SUB_021 | 70's | M | 14 | 14 | 2 | 18 | 2 | 12 | 231 | 32 |
| SUB_022 | 70's | M | 16 | 14 | 6 | 25 | 2 | 23 | 188 | 22 |
| SUB_023 | 50's | M | 24 | 24 | 8 | 21 | 6 | 25 | 42 | 19 |
| SUB_024 | 70's | M | 26 | 21 | 5 | 20 | 10 | 27 | 181 | 19 |
| SUB_025 | 60's | F | 14 | 21 | 7 | 24 | 7 | 26 | 76 | 16 |
| SUB_026 | 60's | F | 20 | 15 | 5 | 23 | 5 | 22 | 114 | 14 |
| SUB_027 | 60's | F | 19 | 8 | 2 | 5 | 3 | 5 | 96 | 31 |

Here , MRgFUS = magnetic resonance guided focused ultrasound, T0 = before MRgFUS, T2 = 1 month after MRgFUS, T3 = 6 months after MRgFUS, M= male, F= female, T= Treated and NT= non-treated side.

**Figure 1.**
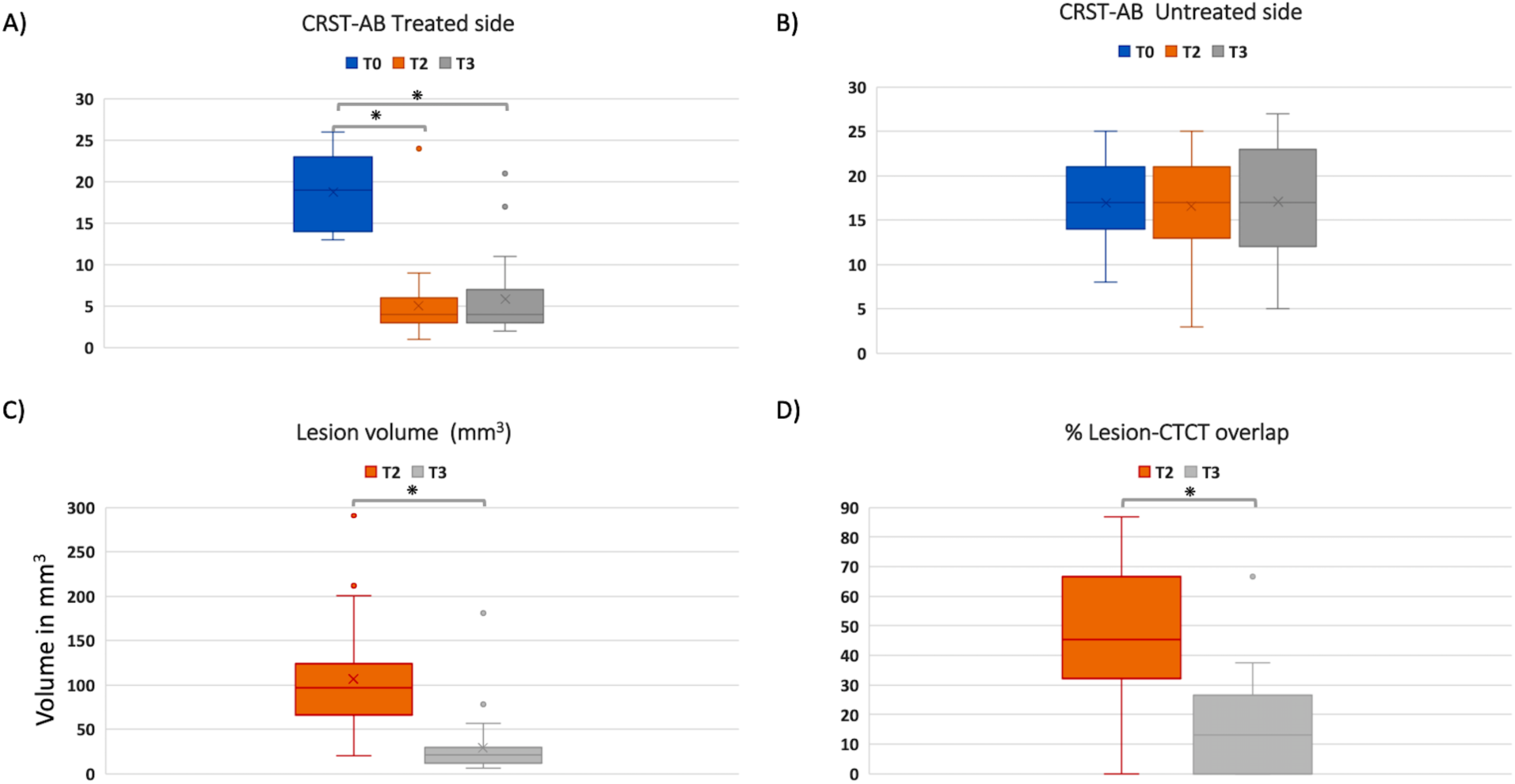
Tremor scores for both, the treated (A) and untreated side (B), before (T0), one month (T2), and six months (T3) following MRgFUS thalamotomy; C) Lesion volume and (D) lesion-CTCT overlap at T2 and T3 following MRgFUS thalamotomy. *p<0.001.

### Lesion volume segmentations

Pairwise comparisons showed a significantly (t(26)=8.39, p<0.001, d= 1.61) reduced lesion volume at T3 (M±SD = 30.9 ± 33.9 mm^3^) compared to T2 (M±SD = 109 ± 62.8 mm^3^) (Figure 1C).

### Lesion-CTCT overlap

We observed 46.5% (range: 0-87%) lesion-CTCT overlap at T2 which dropped to 16.1% (0-66. 7%) at T3 (t(26)=7.50, p<0.001, d= 1.44).

### Microstructural changes within the CTCT

LME models showed significant effects of time at the thalamic lesion site for FA (p<0.001, α<0.05, k=132), AD (p<0.001, α<0.05, k=909), and MD (p<0.001, α<0.05, k=335) (Figure 2: Supplementary Figure 1 for corresponding box plots), but not RD. Post-hoc comparisons at T2 showed a significant decrease in FA (p<0.001, α<0.0166, k=196) and AD (p<0.001, α<0.0166, k=723) at the lesion site compared to T0 baseline (Figure 2). Meanwhile, no significant decreases relative to T0 were observed at T3. Instead, AD (p<0.001, α<0.0166, k=1250) and MD (p<0.001, α<0.0166, k=865) increased significantly at T3 compared to T2.

**Figure 2.**
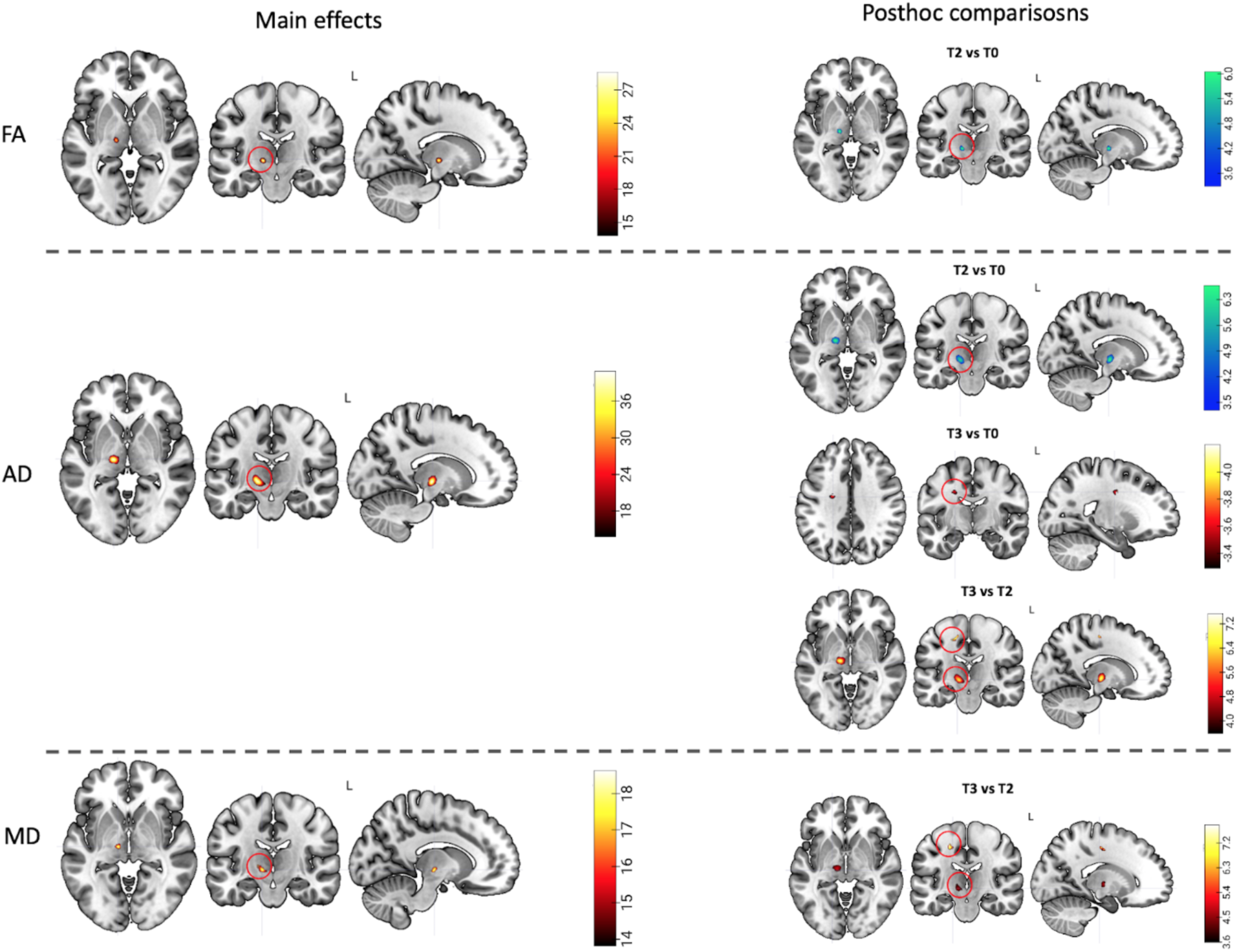
Microstructural changes in FA, AD and MD over time. Left: Local and distant main effects within the treated CTCT (cluster-level α<0.05 for main effects, with cluster-defining height threshold p<0.001 voxelwise). Right: Post-hoc contrasts corrected for multiple comparisons (α<0.0166: 0.05/3). To increase the visibility, we have highlighted (red circles) the significant clusters in the coronal plane. Color bars represent chi-square values for the ANOVA main effect and t-values for post-hoc comparisons (red-hot colors represents increases, blue-cool colors represent decreases). T0 = before MRgFUS, T2 = 1 month following MRgFUS, T3 = 6 months following MRgFUS, FA= fractional anisotropy, AD= axial diffusivity, and MD= mean diffusivity.

Similar increases of AD (p<0.001, α<0.0166, k=170) and MD (p<0.001, α<0.0166, k=184) for T3 relative to T2 were also found in a distant part of the CTCT between thalamus and precentral gyrus (p<0.016) (Figure 2): For the respective comparison versus T0, the same region also showed a cluster (k=132) with increased AD values at T3 (Figure 2) which just missed the cluster extent threshold (p<0.001, α<0.0166, k≥133).

Within the CTCT of the untreated hemisphere, no significant time effect was found for any of the DWI metrics.

### Correlations

Higher overall lesion volume at T2 showed only a trend-level positive correlation (τ=0.244, p=0.076) with stronger short-term tremor reduction at T2, as expressed by percent change in CRST-AB_(T0-T2)/T0_ (Figure 3A), but not at with tremor reduction at T3 (CRST-AB_(T0-T3)/T0_) (τ=0.14, p=0.31). Lesion-CTCT overlap at T2 showed a marginally significant positive correlation with CRST-AB_(T0-T2)/T0_ (τ=0.26, p=0.06), and significantly with long-term percent change in CRST-AB_(T0-T3)/T0_ (τ=0.331, p=0.017, Figure 3B). Neither lesion size nor lesion-CTCT overlap at T3 correlated with the clinical tremor improvement.

**Figure 3.**
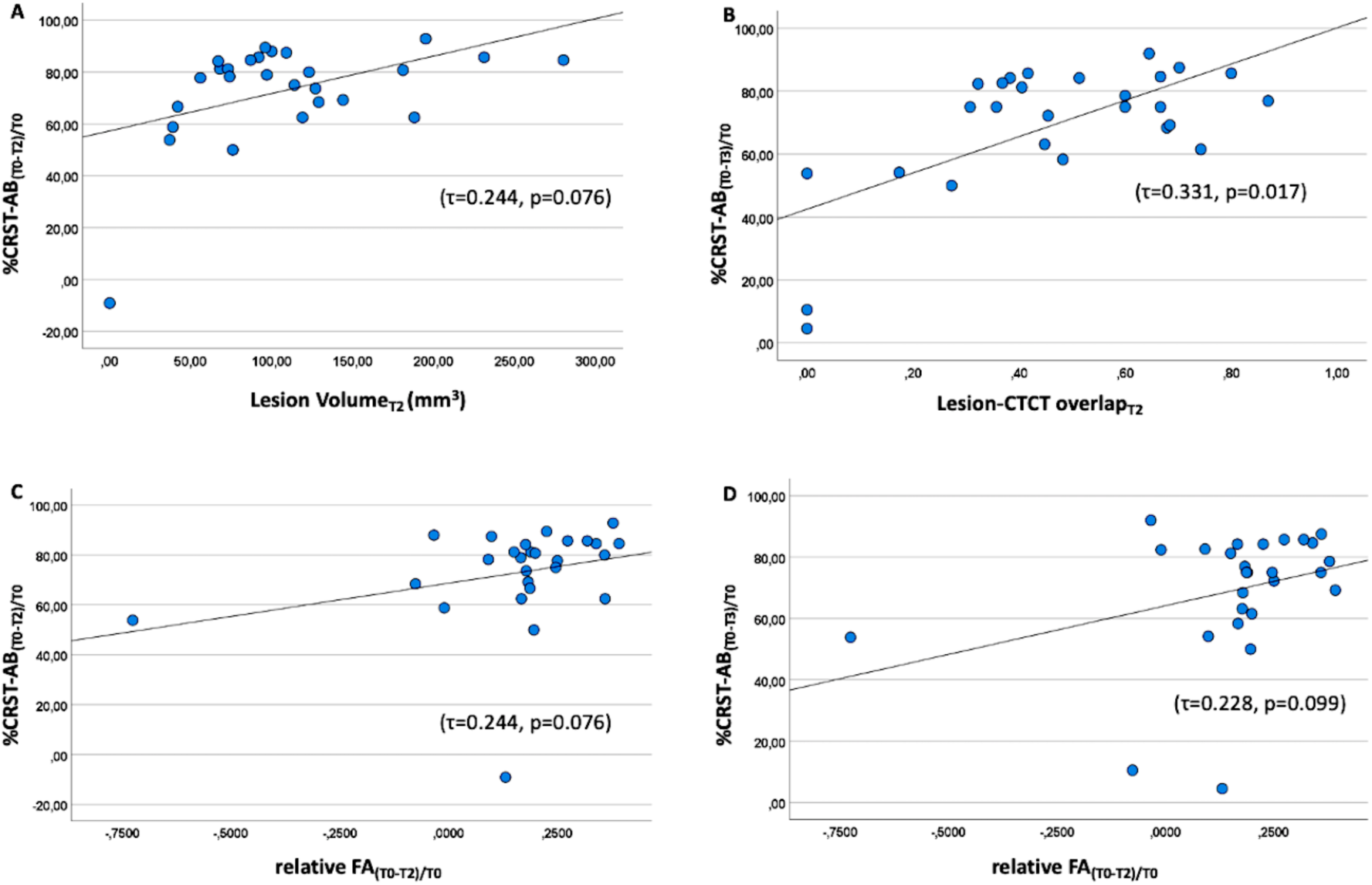
Relationship between: A) lesion volume at 1 month (T2) correlates with percent CRST-AB, B) lesion-CTCT overlap at 1 month correlates with percent CRST-AB at 6 months (T3); relative FA changes at 1 month correlate with C) percent CRST-AB at 1 month and D) percent CRST-AB at 6 months. τ represents the Kendall’s rank correlation with the corresponding p-value. T0 = before MRgFUS, T2 = 1 month following MRgFUS, T3 = 6 months following MRgFUS, FA = fractional anisotropy, CTCT = cerebello-thalamo-cortical tract and CRST = Clinical Rating Scale for Tremor.

Regarding the significant clusters derived for the main DWI analyses, only short-term change in relative FA_(T0-T2)/T0_ for the thalamic cluster showed trend-level positive correlations with the short-term percent change in CRST-AB_(T0-T2)/T0_ (τ=0.244, p=0.076, Figure 3C) and long-term percent change in CRST-AB_(T0-T3)/T0_ (τ=0.228, p=0.099, Figure 3D). No significant correlations were observed between the AD and MD changes for the distant CTCT cluster and clinical tremor improvement.

## DISCUSSION

To our knowledge, this is the first MRgFUS study in patients with severe pharmacoresistant ET which investigated longitudinal voxel-wise changes of microstructural integrity along the treated CTCT fiber tract beyond the first three months of intervention. Examining both short-(one month) and longer-term (six months after treatment) effects, the study provided further insights into the dynamics of the microstructural changes after MRgFUS. Apart from a significant and sustained reduction of tremor severity, and a strong decline in macroscopic lesion volume, microstructural changes in FA, AD, and MD were primarily observed at the lesion site, but also in a distant CTCT section of the treated hemisphere. These microstructural changes showed differential temporal dynamics over the 6-month time course: While FA at the thalamic lesion site showed a decrease at one month, but not at six months after MRgFUS treatment, AD presented an initial decrease followed by a subsequent increase after six months, which was paralleled by MD increases emerging after six months. Findings suggest some restitution of fiber organization in the damaged region, but no full recovery back to the original pre-intervention state. On the other hand, both AD and MD showed a delayed increase at six months (compared to one month after MRgFUS) in an upper aspect of the CTCT (between thalamus and precentral gyrus), which suggests longer-term fiber changes beyond the original site of injury, possible due to chronic degeneration. Meanwhile, reductions in tremor severity were not associated with these distant changes, but only with lesion-specific imaging measures from the 1-month follow-up: Overall, the most robust association with long-term tremor reduction was observed between the percent overlap of the macroscopic lesion with the CTCT tract, even though FA changes showed a similar trend for both, short- and long-term outcomes.

The stable reductions in tremor severity at one and six months after unilateral MRgFUS treatment align with several recent studies (Wintermark, Huss, et al., 2014; Pineda-Pardo et al., 2019; Hori et al., 2020; Zur et al., 2020; Kapadia et al., 2020; Purrer et al., 2021; Mazerolle et al., 2021; Thaler et al., 2023). Yet, the physiological substrates of these therapeutic effects, especially post-acute and long-term structural and functional adaptations of the involved brain networks help to sustain tremor reduction, are only partially understood. Considering the ablation itself, lesion characteristics such as size and anatomical overlap with white matter tracts have been reported to relate to improvement in clinical tremor (Miller et al., 2019; Kapadia et al., 2020; Purrer et al., 2021; Kyle et al., 2023). Similarly, we observed that after one month overall lesion volume marginally correlated with concurrent improvement in tremor severity, but it was not associated with tremor change after six months. Instead, a lesion-CTCT overlap metric was significantly correlated with long-term tremor improvement further validating the previous findings that the degree by which the lesion affects the CTCT is predicting long-term tremor improvement in ET.

Microstructural integrity can be altered due to pathology or treatment and characterized using diffusion measures (Alexander et al., 2007). Previous studies in ET patients have reported a decrease in FA locally at lesion location as well as remotely in cerebellar peduncle and between thalamus and motor cortex at longer-term following MRgFUS (Pineda-Pardo et al., 2019; Mazerolle et al., 2021; Thaler et al., 2023) suggesting that loss or increased incoherency of fibers leads to reduced microstructural integrity (Walker et al., 2019). In line with these, we observed significant decreases in thalamic FA at one month following MRgFUS treatment, which at six months showed some evidence for recovery, as no significant difference from baseline was found here (Supplementary Figure 1). Earlier study had reported significant FA decreases at three months (Wintermark, Huss, et al., 2014), and there is some evidence for incomplete recovery (Zur et al., 2020). Meanwhile, Thaler et al. (2023) reported FA decreases even after one year, but this could be due to larger residual lesions size (mean: 42.28 ± 23.12 mm^3^) after one year as compared to our study at only six months (30.9 ± 33.9 mm^3^) which might reflect more extensive and sustained tissue loss in their patients. Concomitant with the FA changes, we observed a differential development of AD changes, an initial decrease at one month and an increase at six months. AD is thought to provide specific information about axonal injury or coherence in the absence of edema (Winston et al., 2014). The short-term local decrease in AD is consistent with an animal study reporting an acute decrease in AD after MRgFUS due to axonal damage (Walker et al., 2019). A residual influence of vasogenic edema should, however, not be discounted, also considering that AD levels increased to baseline levels at six months follow-up. Meanwhile, MD showed a relative increase from one month to six months, which may reflect chronic inflammatory processes (Oestreich & O’Sullivan, 2022).

Unlike previous studies (Pineda-Pardo et al., 2019; Mazerolle et al., 2021; Thaler et al., 2023), we did not observe remote FA changes. However, we obtained delayed increases in AD and MD in a distant region of the CTCT located below the motor cortex at six months, mainly compared to measurements at one month. These findings extend prior observations of AD or MD increase following three months (Mazerolle et al., 2021) and even at one year (Thaler et al., 2023) after MRgFUS. MRgFUS lesion leads to axonal damage (Walker et al., 2019), triggering a cascade of cellular and metabolic responses leading to Wallerian degeneration distal to the injury site (Conforti et al., 2014). Accordingly, the remote MD increases found in our data and these previous studies would be consistent with this sort of protracted fiber loss, as increased MD is frequently interpreted to reflect reduced white matter integrity due to either axonal or myelin degradation (Solowij et al., 2017). Meanwhile, the colocalized increase of AD which was observed both in our data and Mazerolle et al (2021) would be more suggestive of regenerative processes, as higher AD would be interpreted to reflect higher axonal caliber or stronger coherent orientation of axons (Winklewski et al., 2018). Given the ongoing debate about the specificity of these measures in identifying pathology, this cannot be clarified (Jones et al., 2013). More generally, the distant changes showed no relationship with tremor reduction, thus, questioning the functional relevance of these distant AD and MD changes for the present clinical outcome.

Previous work has shown lesion FA values to correlate with tremor outcomes (Hori et al., 2020; Zur et al., 2020). We only observed marginally significant associations between voxel-wise thalamic FA decrease and tremor reduction at short and long-term following MRgFUS. There were also other studies that did not find significant associations between lesion-specific DWI metrics and clinical outcomes (Pineda-Pardo et al., 2019). Methodological factors which vary between studies, including sample size, lesion volume, lesion-tract overlap, or variance of clinical outcomes, may contribute to the variability of these associations.

This study is not without limitations. The moderate sample size of this study limits generalizability and statistical power. Nevertheless, we were able to find significant microstructural changes in the targeted CTCT. The parameters of the DWI acquisition could be improved to attain higher spatial resolution, e.g., by using multi-shell acquisitions, in order to detect more subtle changes. We used a tractography-guided lesion approach, which accounts for individual anatomical variation, but may result in greater variability in lesion localization than an independent atlas-based method. By extending the number of longitudinal MR examinations, a more reliable estimation of the temporal trajectories of the observed changes would become possible. Finally, the inclusion of clinical tremor scores from longer-term follow-ups could better distinguish between patients with stable outcomes, and future symptom recurrences (Kapadia et al., 2020). Moreover, clinical characterization could be augmented by objective measures (e.g., accelerometry).

## Conclusion

This study further substantiates both short-term and long-term microstructural changes following unilateral MRgFUS in ET patients, as reported in prior studies with smaller cohorts, and often shorter follow-ups. Beyond temporary reductions of diffusion MRI measures at the lesion site which were recurrent, and indicate axonal disruption or incoherency after MRgFUS in the short term, potentially followed by axonal reorganization in the longer term, the present study also confirmed delayed microstructural changes in upper aspects of the CTCT which may reflect remote lesion effects on CTCT integrity, e.g., due to Wallerian-like degeneration. This warrants further validation with refined imaging protocols, especially to improve their limited potential as a predictive marker for treatment outcomes.

## Data Availability

All Data produced in the study contained patient related information which is not available publicly.

## Abbreviations

MRgFUS: Magnetic Resonance Guided Focused Ultrasound
ET: Essential tremor
VIM: Vental intermediate Neucleus
CTCT: cerebello-thalamo-cortical tract
T0: Before MRgFUS
T2: 1 month after MRgFUS
T3: 6 months after MRgFUS
CRST: Clinical rating scores for tremor
DWI: Diffusion Weighted Imaging
FA: Fractional Anisotropy
AD: Axial Diffusivity
MD: Mean Diffusivity

## Acknowledgment

We acknowledge all the participants and their relatives for their support in carrying out this study. We also acknowledge the medical radiology assistants who performed the clinical data acquisition as well as the neurologists involved in clinical assessments.

## Author contributions

The MRgFUS study was conceptualised and designed by Henning Boecker and Ullrich Wüllner. Resources were largely provided by Carsten Schmeel, Alexander Radbruch, and Henning Boecker. Ullrich Wüllner, Veronika Purrer and Valeri Borger were involved in the MRgFUS treatment and other clinical aspects. The data analysis was carried out by Neeraj Upadhyay. Data curation was done by Neeraj Upadhyay, Veronika Purrer, and Jonas Krauß. Statistical analysis and interpretation of the data was carried out by Neeraj Upadhyay, Marcel Damen, and Henning Boecker. Neeraj Upadhyay, Angelika Maurer, Marcel Damen and Henning Boecker drafted and edited the manuscript. All authors critically reviewed and edited the manuscript and approved the final manuscript.

