## Supplementary Figure 1 for "Longitudinal Evolution of Diffusion Metrices in the Cerebello-Thalamo-Cortical Tract After MRgFUS Thalamotomy for Essential Tremor"

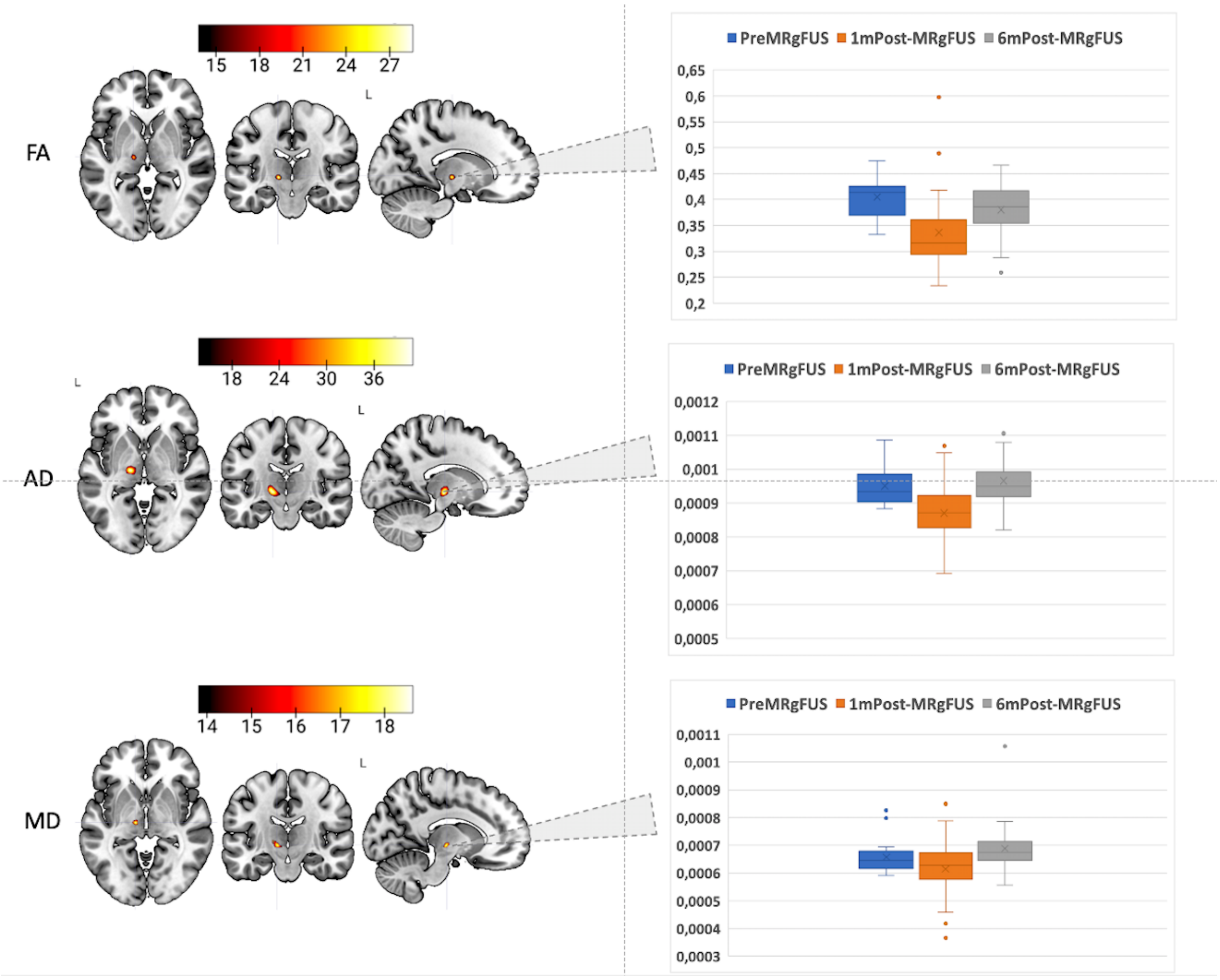


Above Figure represent the bar plots for FA, MD and AD created from the main time effect clusters over the time PreMRgFUS (T0), 1m Post-MRgFUS (T2), and 6mPost-MRgFUS (T3).
